# Importance of epidemic severity and vaccine mode of action and availability for delaying the second vaccine dose

**DOI:** 10.1101/2021.06.30.21259752

**Authors:** Luděk Berec, René Levínský, Jakub Weiner, Martin Šmíd, Roman Neruda, Petra Vidnerová, Gabriela Suchopárová

**Affiliations:** Centre for Mathematical Biology, Department of Mathematics, Faculty of Science, University of South Bohemia, Branišovská 1760, 37005 České Budějovice, Czech Republic; The Czech Academy of Sciences, Biology Centre, Institute of Entomology, Department of Ecology, Branišovská 31, 37005 České Budějovice, Czech Republic; CERGE-EI, Politických vězňů 7, 11121 Praha 1, Czech Republic; Siesta Labs, Konopišťská 16, 10000 Praha 10, Czech Republic; The Czech Academy of Sciences, Institute of Information Theory and Automation, Pod Vodárenskou věží 4, 18200 Praha 8, Czech Republic; The Czech Academy of Sciences, Institute of Computer Science, Pod Vodárenskou věží 2, 18200 Praha 8, Czech Republic

## Abstract

Following initial optimism regarding the potential for rapid vaccination, delays and shortages in vaccine supplies have occurred in many countries. Various strategies to counter this gloomy reality and speed up vaccination have been set forth, of which the most popular approach has been to delay the second vaccine dose for a longer period than originally recommended by the manufacturers. Controversy has surrounded this strategy, and overly simplistic models have been developed to shed light on this issue. Here we use three different epidemic models, all accounting for the actual COVID-19 epidemic in the Czech Republic, including the rise and eventual prevalence of the B.1.1.7 variant of SARS-CoV-2 virus and real vaccination rollout strategy, to explore when delaying the second vaccine dose from 21 days to 42 days is advantageous. Using the numbers of COVID-19-related deaths as a quantity for comparing various model scenarios, we find that vaccine mode of action at the beginning of the infection course (preventing contagion and symptom appearance), mild epidemic and sufficient vaccine supply rate call for the original inter-delay scenario of 21 days regardless of vaccine efficacy. On the contrary, for vaccine mode of action at the end of infection course (preventing severe symptoms and death), severe epidemic and low vaccine supply rate, the 42-day inter-dose period is preferable, at any plausible vaccine efficacy.

**One sentence summary:** We address when delaying the second vaccine dose is advantageous, considering various epidemic severities and various vaccine actions and availabilities.

## Introduction

Although recent modeling studies cast some doubt on earlier optimism that vaccination may mean an end to the COVID-19 pandemic (*1,2*), it undoubtedly provides significant leverage that at least temporarily may bring back a return to close-to-normal life. Even so, while everything sounded promising during the autumn 2020 months, delays and shortages in vaccine supplies in spring 2021 led to complications and spurred thinking on strategies that would speed up vaccination, despite limited vaccine supplies. Indeed, models have clearly demonstrated that vaccine deployment delays substantially affected the course of the pandemic (*3, 4*). Eventually, the strategy of delaying the second dose, required for all actually available vaccines except the Janssen one produced by Johnson & Johnson (*5*), has often been adopted.

This has also been the case in the Czech Republic (also known as Czechia). As of May 8, 2021, over 3.5 million vaccine doses were applied (about 2.5 million first doses and about 1 million second doses), of which nearly 79% were BNT162b2 (Pfizer/BioNTech) and nearly 10% Moderna (*6*). Until April 1, 2021, the second dose was delayed after the first one as recommended by the vaccine producers (21 days for BNT162b2 and 28 days for Moderna (*7*)). Since then, the government allowed the increasing of the second dose delay for both vaccines to six weeks, which is now the standard vaccination scheme in Czechia (*8*).

As with many other issues concerning the COVID-19 pandemic, models have been developed to guide formulation of vaccination strategies. The first set of models of this kind focused on how to distribute a limited number of vaccines within the population; that is, which groups of people to prioritize. With the objective to avert as many deaths as possible, a unanimous answer to this question has been to start with vaccinating the oldest age cohorts and then continue by gradually including younger age cohorts, as a consequence of an accelerating age-dependent infection mortality profile (*9, 10*). This strategy has been adopted by virtually every country. An obvious exception to this has been a preferential vaccination of the health workers directly interacting with COVID-19 patients.

Given the limited supply of vaccines, the strategy of vaccinating more people by delaying the second dose is at a glance natural. Indeed, the trade-off appears to be between vaccinating a given number of individuals with just one dose, despite reduced efficacy and in the hope that more vaccines will arrive shortly, as opposed to vaccinating half that number and holding one dose back for each administered first dose. Accounting for different protection levels reached after the first and second doses, existing modeling studies suggest that such a “dose-sparing” strategy should indeed be considered, as it would avert more cases of COVID-19 and more deaths due to COVID-19 compared with when an original scheme is used (*11, 12*).

However, the existing models are too simple to give us robust direction regarding how to proceed. They either do not account for epidemic dynamics (*12*) or choose between a one-dose scheme and the recommended two-dose scheme administered to half the people, using for many countries currently quite unrealistic values of the effective reproduction number (*R* = 1.8*−*2.1) (*11*). In parallel with vaccination, an epidemic continues and many people still get infected and eventually immunized, or die. Moreover, the vaccine supply and distribution are themselves is dynamic: in the two-dose scheme, we do not set aside one vaccine per any applied one, because others are coming on a more or less regular basis. None of the studies thus appears to model any current epidemic and any more or less realistic vaccine rollout scenario.

Here we explore whether and when delaying the second vaccine dose from 21 days originally suggested for the BNT162b2 vaccine (Pfizer/BioNTech) by additional three weeks to 42 days is advantageous, on the basis of the numbers of COVID-19-related deaths averted by June 30, 2021. In doing this, so as to cover many plausible situations, we consider a variety of potential vaccine efficacies, modes of vaccine action, epidemic severities, and (time-dependent) vaccine supply rates. In addition, to provide robust results, we use three different epidemic models that all independently account for real epidemic in Czechia, including the rise and eventual prevalence of the B.1.1.7 variant of SARS-CoV-2 virus early in 2021, and the actual vaccination rollout strategy. All three models are of the SEIR type, the generally agreed framework to model COVID-19, and all explicitly consider both the asymptomatic and presymptomatic infectious classes. That is, once infected, individuals remain non-infectious for a (latent) period. Then, a proportion of them become infectious yet asymptomatic for the rest of infection, while the others become symptomatic following a short presymptomatic (yet already infectious) period. Symptomatic individuals either recover or die, with different models using various other states of infectiousness within this period. Moreover, all models are structured by age of individuals and type of inter-individual contacts. Nonetheless, each model also has a number of unique assumptions and characteristics (Methods).

## Results

In Czechia, and also in many other countries, the BNT162b2 (Pfizer/BioNTech) vaccine is the dominat one. We thus motivate timing of individual vaccine doses by this vaccine brand. The first dose efficacy is commonly reported to establish roughly two weeks after its application, while the second dose efficacy is fully unrolled about one week after its application (Table 1). Also, we assume that the first dose efficacy holds until one week after the second dose application, not considering any vaccine efficacy fadeout before the second dose. These assumptions are common in modeling studies (*2, 11*), although other assumptions can also be made (*12*).

**Table 1:** Published values on efficacy for the Comirnaty (Pfizer/BioNTech) vaccine. Modes of action: H - preventing hospitalization, T - preventing transmission, S - reducing probability of symptoms appearance upon infection, J - preventing severe or critical state in hospitals, D - preventing COVID-19 related death. The study of (*19*) also shows only negligible dependence on age and further slight increase in vaccine efficiency 14 days after the second dose.

| 1st dose VE<br>(% (95% CI)) | 2nd dose VE<br>(% (95% CI)) | Mode of<br>action | Remark | Source |
| --- | --- | --- | --- | --- |
| 52.4 (29.5-68.4) | 94.8 (89.8-97.6) | ? | Early assessment by Pfizer, after 1st dose and 7 days after 2nd dose | (14) |
| 85 (76-91) |  | H | Increasing over time, peaking at 28-34 days post-vaccination | (15) |
| 72 (58-86) | 86 (76-97) | T&S (?) | 21 days after 1st dose and 7 days after 2nd dose, dominant variant B.1.1.7 | (16) |
| 68.5 (46.5-81.5) |  | ? | 7 days after the 1st dose, criticism of (14) | (17) |
| 92.6 (69-98.3) |  | ? | 14 days after the 1st dose, criticism of (14) | (17) |
| 46 (40-51) | 92 (88-95) | T | 14-20 days after first dose, 7 days after 2nd dose | (18) |
| 57 (50-63) | 94 (87-98) | S | 14-20 days after first dose, 7 days after 2nd dose | (18) |
| 74 (56-86) | 87 (55-100) | H | 14-20 days after first dose, 7 days after 2nd dose | (18) |
| 62 (39-80) | 92 (75-100) | J | 14-20 days after first dose, 7 days after 2nd dose | (18) |
| 72 (19-100) |  | D | 14-20 days after first dose | (18) |
|  | 95.3 (94.9-95.7) | T | 7 days after 2nd dose | (19) |
|  | 97 (96.7-97.2) | S | 7 days after 2nd dose | (19) |
|  | 97.2 (96.8-97.5) | H | 7 days after 2nd dose | (19) |
|  | 97.5 (97.1-97.8) | J | 7 days after 2nd dose | (19) |
|  | 96.7 (96-97.3) | D | 7 days after 2nd dose | (19) |

Since our major question is whether and when delaying the second vaccine dose beyond the period recommended by producers is advantageous, we assume that the second dose is applied either 21 or 42 days after the first one and present the results following a uniform structure: given a model of vaccine action (or a combination of model), vaccine supply rate and epidemic severity, then for any plausible vaccine efficacy combination we, as our main summary statistic, calculate the amount of COVID-19-related deaths averted among adults 65+ years by June 30, 2021, when adopting the longer delay. To account for inevitable uncertainty in model predictions, each model is run for repeatedly (Models M and F) or over different plausible parameter sets (Model H; Methods) and the average summary statistic is provided. Whereas blue shades in the figures that follow indicate that 21-day inter-dose period is preferable, in the red shaded regions more deaths are averted when 42-day inter-dose period is used.

### Effects of vaccine action

We consider a number of plausible vaccine actions; see Methods for their detailed description. The mode of vaccine action has a non-negligible effect on whether it is advantageous to delay the second dose by 42 rather than 21 days (Figs 1-3). In particular, delaying the second vaccine dose by 42 rather than 21 days is most beneficial and significant when the vaccine acts simultaneously on the probabilities of needing an ICU when in hospital and dying when in the ICU (Fig. 3 bottom right). On the contrary, the effect seems virtually negligible when the vaccine acts simultaneously on the earliest two elements in the infection progression (probabilities of (leaky) transmission and becoming symptomatic; Fig. 1 bottom row).

**Figure 1:**
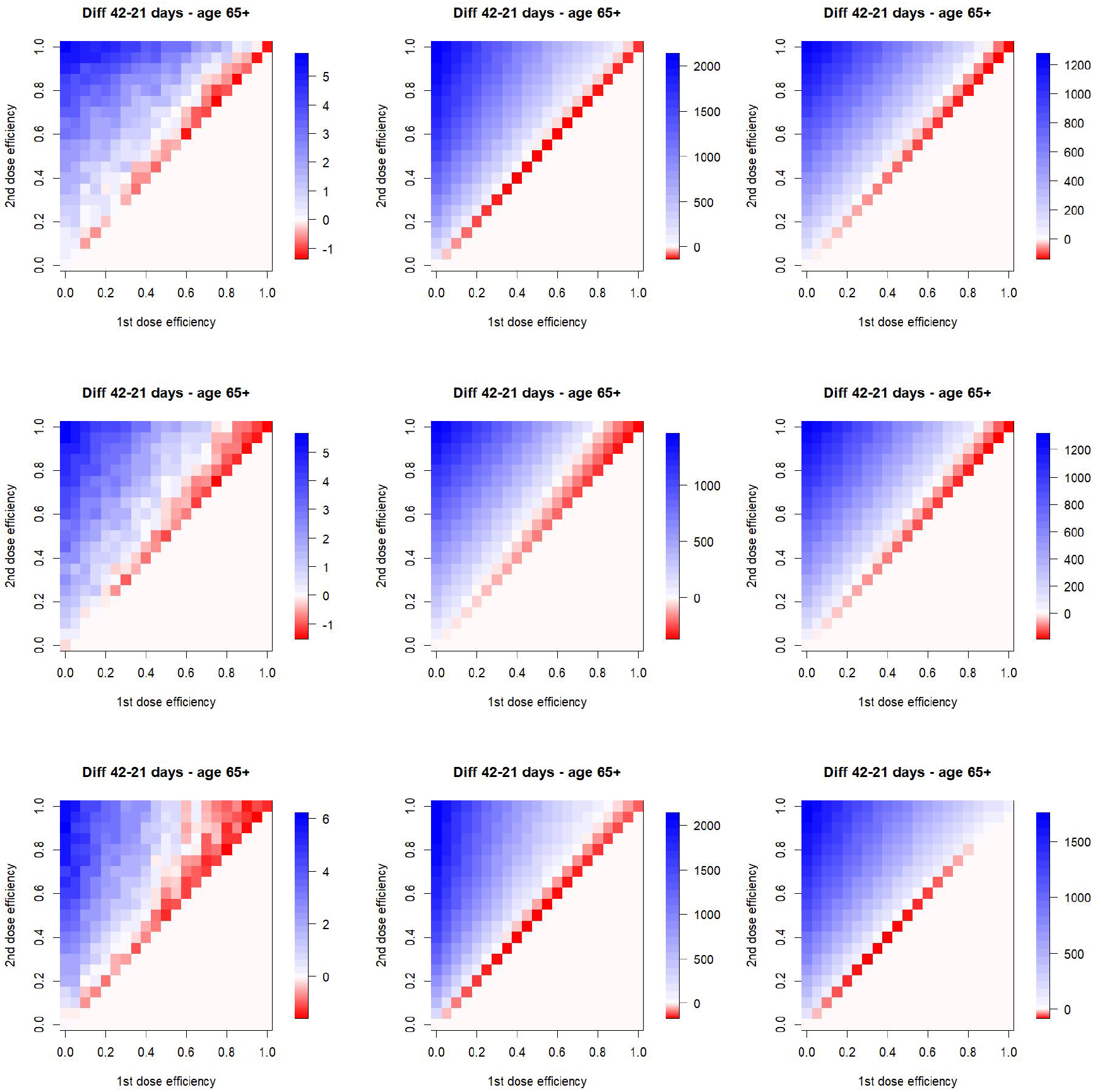
Efficacy of delaying the second vaccine dose by three weeks from 21 to 42 days, for different modes of vaccine action and different adopted epidemiological models. Top row: leaky vaccine effect on the probability of infection transmission, second row: vaccine effect on the probability of becoming symptomatic when infected, bottom row: combination of leaky transmission and probability of becoming symptomatic. Left column: model M, middle column: model H, right column: model F.

**Figure 2:**
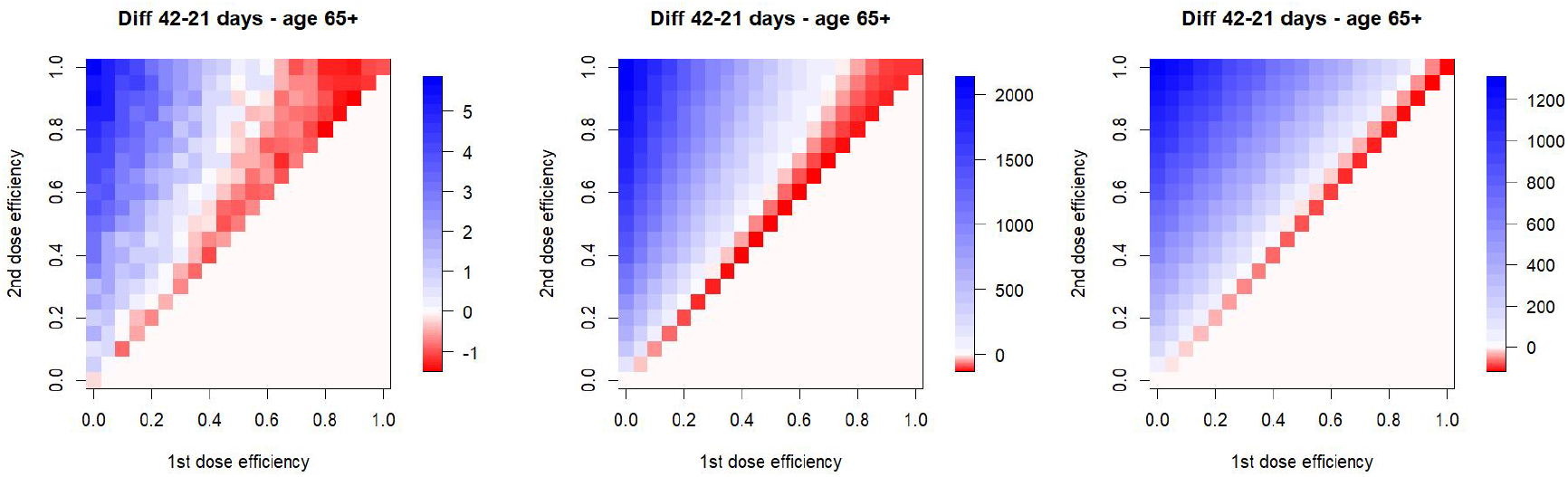
Efficacy of delaying the second vaccine dose by three weeks from 21 to 42 days, for all-or-nothing vaccine effect on the probability of infection transmission. Left column: model M, middle column: model H, right column: model F.

Importantly, effects of vaccine actions in different steps of infection progression do not add up but rather multiply. This of course follows trivially from the model formulation, but may be surprising at first glance. Just compare the vaccine effect o leaky transmission (Fig. 1 top row), probability of symptoms appearance (Fig. 1 middle row) and their combined effect (Fig. 1 bottom row). Or alternatively the effect on hospitalization probability (Fig. 3 left), probability of symptom appearance (Fig. 1 middle row), and on all elements during infection progression (Fig. 3 right). The multiplicity of the modes of action means that when the infection of a proportion of individuals in a model state are averted due to a vaccine action, only a proportion of the infections in remaining proportion are averted in a following state of infection progression (symptoms appearance, probability of getting hospitalized, needing an ICU, or dying).

**Figure 3:**
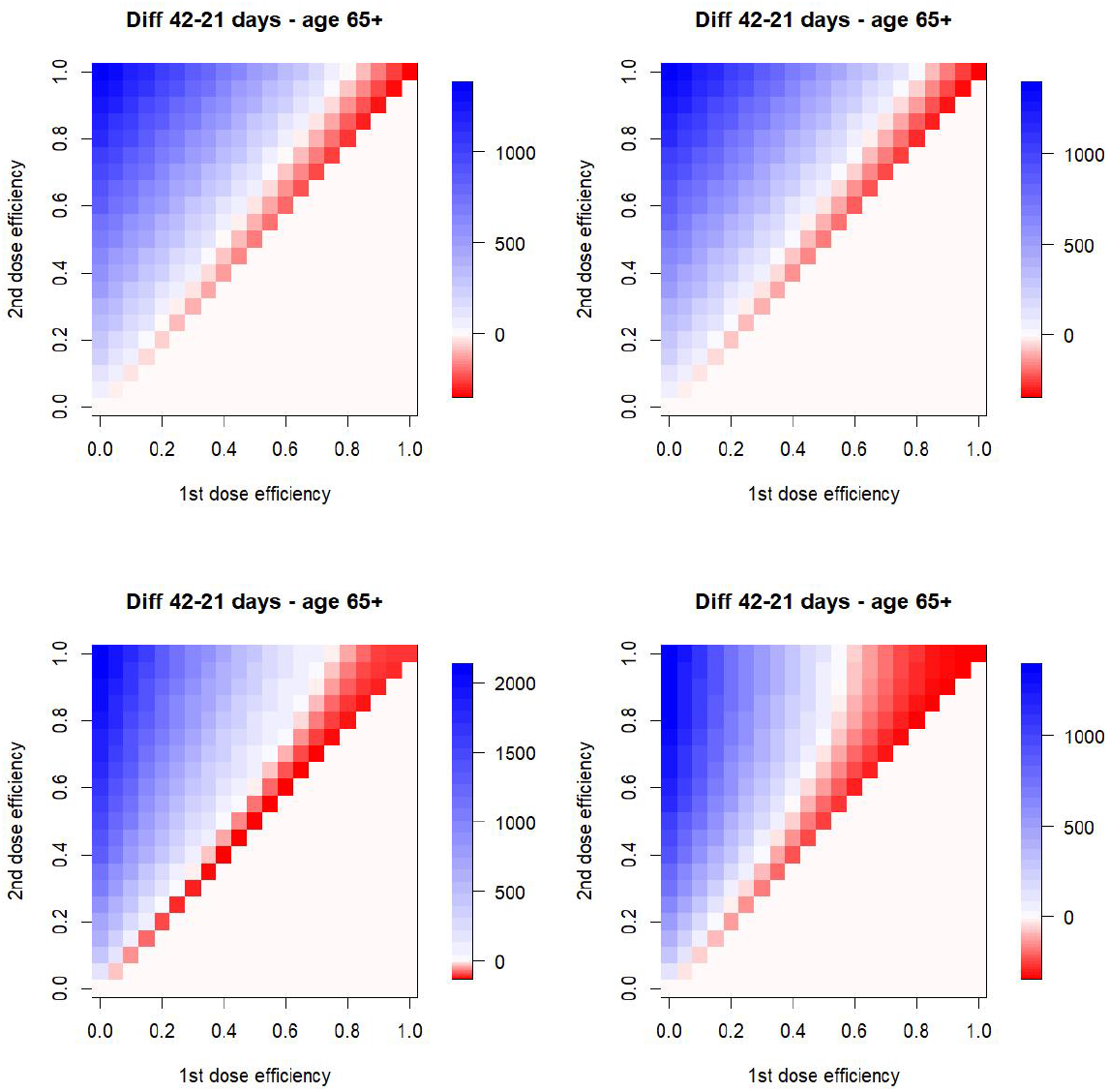
Efficacy of delaying the second vaccine dose by three weeks from 21 to 42 days, assuming vaccine effect on the probability that a symptomatic individual becomes hospitalized (top left), the probability that a hospitalized individual needs an ICU (top right), the probability of dying when on the ICU (bottom left), and combination of effects on the probability of needing an ICU and probability to die when on ICU (bottom right). Only model H is used here.

### Effects of epidemic severity

The above results are based on the actual infection severity of epidemic in the Czech Republic. This is to a large extent driven by the contact structure, estimated to be at 45% of the prepandemic state since March 1, 2021 (Methods). An increase in the number of contacts, that is, in epidemic severity, corresponding to a uniform increase in the effective reproduction number over time, results in a larger set of vaccine efficacy combinations for which it is advantageous to delay the second dose for 42 days (Fig. 4). Importantly, this effect is analogous for any mode of vaccine action (or their combination; just the combination of effects on infection transmission and symptoms appearance shown here). In conclusion, the more severe the epidemic is, the more advantageous is to delay the second vaccine dose by 42 days.

**Figure 4:**
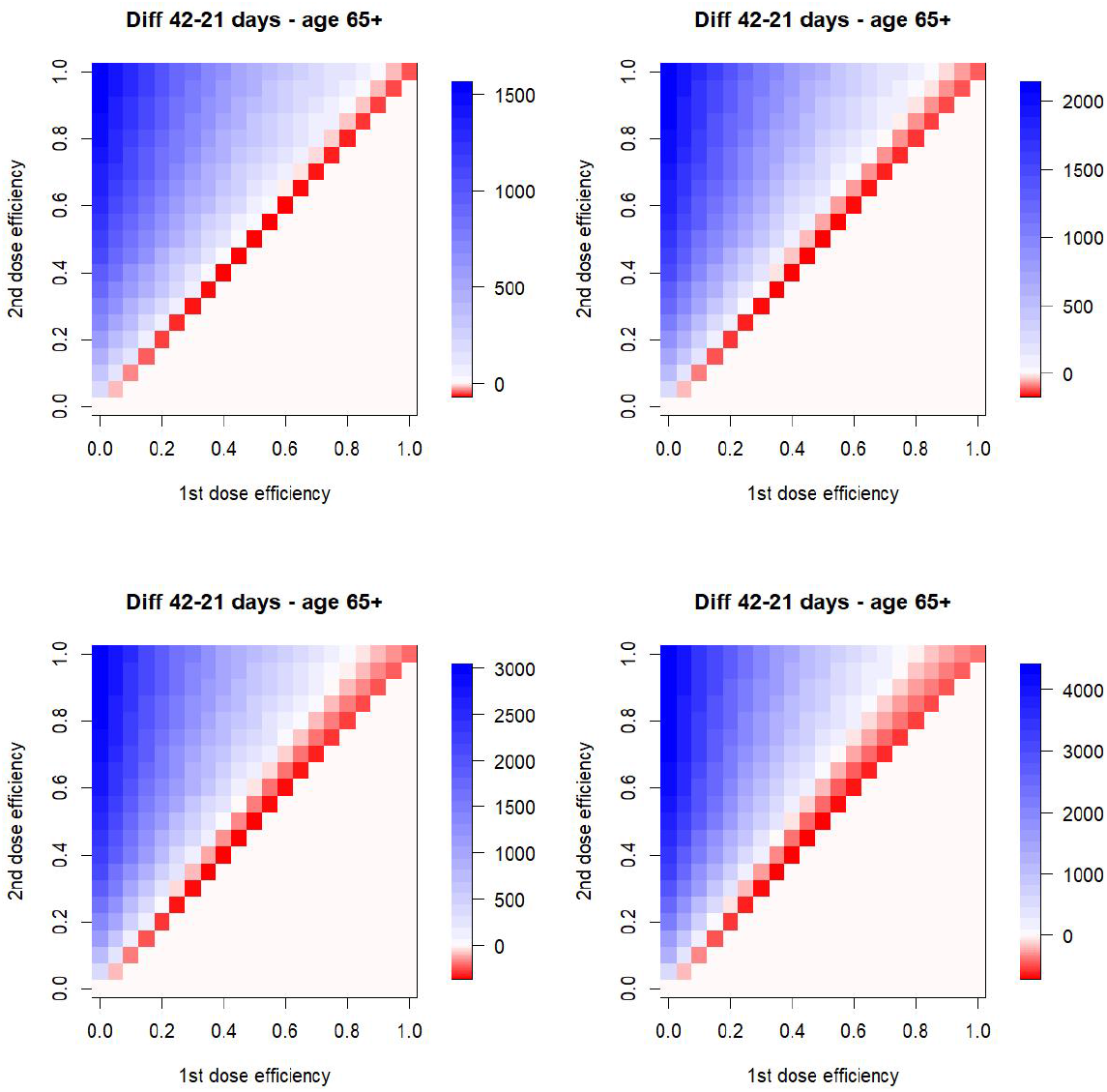
Efficacy of delaying the second vaccine dose by three weeks from 21 to 42 days, assuming leaky vaccine effect on the probability that a vaccinated person gets infected by contact with an infectious one, and at the same time on the probability that a vaccinated person becomes symptomatic when infected. Top left: contacts at 35% of the pre-pandemic state, top right: contacts at 45% (real situation in Czechia), bottom left: contacts at 55%, bottom right: contacts at 65%. Only model H is used here.

### Effects of vaccine availability

We also test effects of vaccine supply scenarios that differ from the actual state in Czechia. As Fig. 5 clearly demonstrates, the less vaccines are available the more advantageous it is to delay the second vaccine dose for 42 days. Again, these results do not change qualitatively if other modes of action (or their combinations) are considered. Interestingly, further improvement in vaccine supply rate relative to the actual vaccination rollout in Czechia does not suggest any significant change in the inter-dose delay preferences, but shortage of supplies, on the other hand, results in much wider vaccine efficacy combinations at which the 42-days delay is advantageous (Fig. 5).

**Figure 5:**
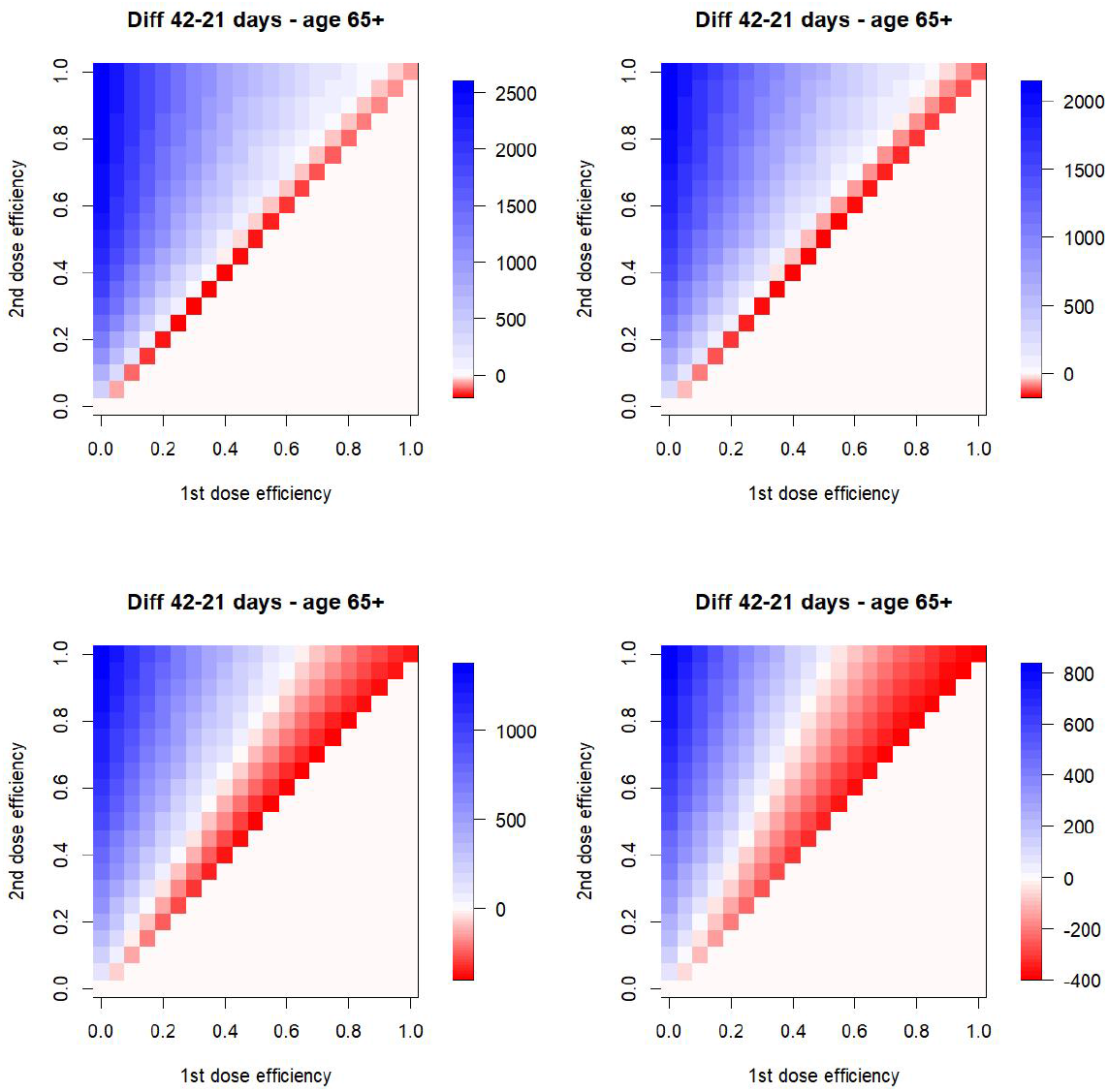
Efficacy of delaying the second vaccine dose by three weeks from 21 to 42 days, assuming leaky vaccine effect on the probability that a vaccinated person gets infected by contact with an infectious one, and at the same time on the probability that a vaccinated person becomes symptomatic when infected. Top left: twice more doses available, top right: actual state in Czechia, bottom left: twice less doses available, bottom right: four time less doses available. Only model H used here.

### The extreme scenarios

Vaccine effect on leaky transmission and probability of having symptoms when infected, mild epidemic and sufficient vaccine supply rate call for the original inter-delay scenario of 21 days regardless of the vaccine efficacy combination (Fig. 6 left). On the contrary, for the vaccine effect on the probabilities of needing an ICU when hospitalized and dying when in the ICU, severe epidemic and insufficient vaccine supply rate, the 42-day inter-dose period is more advantageous than 21 days at any plausible vaccine efficacy combination: if the first dose is more than about 50% efficient, it is beneficial to delay the second dose by other three weeks at any efficacy of the second dose. (Fig. 6 right).

**Figure 6:**
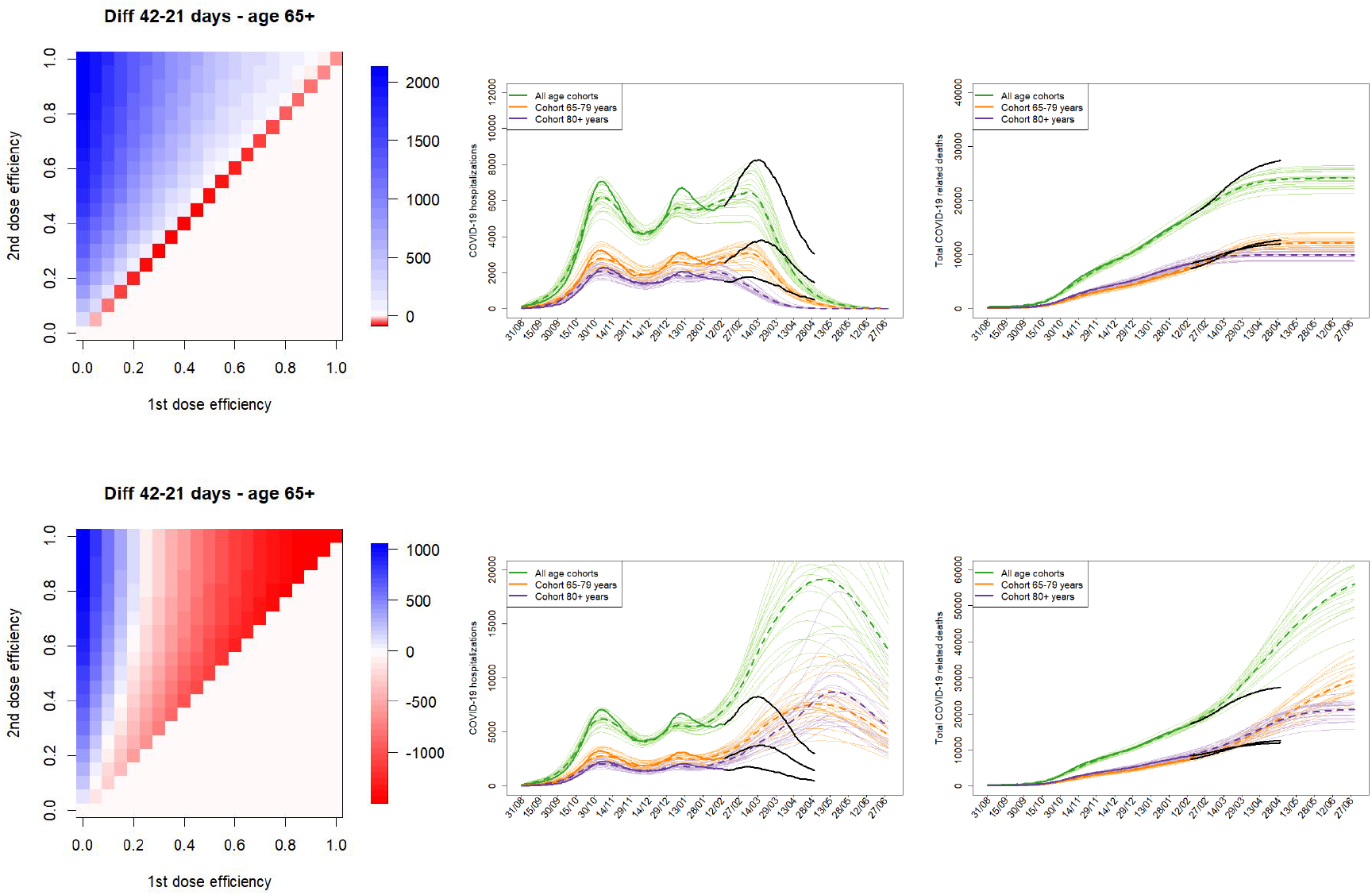
The extreme scenario. Efficacy of delaying the second vaccine dose by three weeks from 21 to 42 days, assuming (top row) vaccine effect on infection transmission and the probability that an infected person shows symptoms, contacts at 35% of pre-pandemic state and twice more doses available, and (bottom row) vaccine effect on the probabilities of needing an ICU and dying when on the ICU, contacts at 65% and four times less doses available. Each of these extreme scenarios is moreover supplemented by the course of the numbers of hospitalized and dead individuals (legend as in Fig. 7 in Methods). Only model H is used here, with 70% and 90% efficacy of the first and second dose, respectively.

## Discussion

The limited supply of COVID-19 vaccines around the world has called for a search for ways to distribute vaccines to a larger number of people. One such strategy, widely used in Europe, is to increase the period between the first and second dose of vaccines based on two-dose schemes. This strategy, based on the idea that it is better to vaccinate more people with just one dose, even if full vaccine efficacy is not reached, rather than to put aside one dose for the re-vaccination for each administered one, is at a glance reasonable. As recently found by a study led by the University of Birmingham, an antibody response in adults 80+ years is 3.5 times larger in individuals that got the second dose of the Pfizer/BioNTech vaccine after 12 weeks compared to those who got it after 21 days (*13*).

The argument that delaying the second vaccine dose is advantageous has received some support also via mathematical models (*11, 12*). Both these studies suggest that such a dose-sparing strategy would avert a significant proportion of infections that would otherwise occur if the originally recommended two-dose vaccination scheme is kept, provided that roughly 50% protection is achieved by the first dose. On the other hand, both studies are simplified in many respects. The study of (*12*) does not account for epidemic dynamics, spans a short time period and as the main comparative statistic considers the numbers of averted COVID-19 cases. The study of (*11*), while based on a SEIR-type epidemic model, does not consider any dynamics in vaccine supply and is based on an artificial epidemic, assuming excessively high effective reproductive numbers (*R* = 1.8 *−* 2.1). Moreover, neither of these studies consider various modes in which this vaccine may act.

Here we attempted to complement these studies and get further insight by: (i) considering a fully dynamic epidemic model calibrated for COVID-19 epidemic in Czechia, (ii) considering actual vaccination rollout scenarios adjusted for actual inter-dose periods and realistic supply rates, (iii) using the number of COVID-19-related death as the comparative statistic, (iv) considering various vaccine efficacies, epidemic severities, and modes in which vaccines may act. Our main conclusion is that vaccine effect on leaky transmission, mild epidemic and sufficient vaccine supply rate call for the original inter-delay scenario of 21 days regardless of the vaccine efficacy combination. On the contrary, for the vaccine effect on probability of dying when on the ICU, severe epidemic and insufficient vaccine supply rate, the 42-day inter-dose period is more advantageous than 21 days at any plausible vaccine efficacy combination. This is currently the case in many Asian and South American countries, as opposed to in many European countries.

In (*12*), the impact of vaccine shortages has a modal form: a moderate decrease in the vaccine supply rate implies more cases averted when the second dose is a bit postponed, but a larger reduction has a clear negative effect and should not lead to postponing the second dose administration. Also, they show that more severe epidemic should mean not prolonging the inter-dose period. Here we show just the opposite: less vaccine availability leads to stronger support for the inter-dose period of 42 days; higher epidemic severity carries the same implication (even though here the effect is not that strong). We also show that postponing the second dose may be more effective even when the first dose efficacy is deeply under 50%, depending on its mode of action, and that the efficiency of the second dose also plays the role.

The vaccine efficacy values published in the literature get more and more precise as new studies arise in response to intense vaccination campaigns in some countries; we provide a summary of those values in Table 1. Overall, it appears that the 14-20 days after first dose the vaccine efficacy level at a value between 50-70%, and 7 days after the second dose the vaccine efficacy settles in between 90-97%, irrespective of the mode of action. We found that the vaccine effect at the beginning of infection progress (transmission, symptom appearance), mild epidemic and sufficient vaccine supply rate call for the original inter-delay scenario of 21 days regardless of vaccine efficacy. On the contrary, for vaccine effect at the end of infection progress (severe symptoms, death), severe epidemic and low vaccine supply rate, 42-day interdose period is better, at any plausible vaccine efficacy.

## Methods

Here we give a short overview of the models we use to address our questions, as well as describe the vaccination rollout scenarios we consider.

### Vaccination scenarios

Two vaccination rollout scenarios are considered, corresponding to delaying the second vaccine dose by either 21 or 42 days. Each scenario provides the daily amount of available first doses (to be) administered to two population groups: general population and health (and other critical infrastructure) workers. Since our models are age-structured (see below), we follow the widely accepted prioritization strategy and in the latter group vaccinate according to age, starting with the eldest individuals. It is important to say that the way epidemic further unrolls depends on its severity and the way the vaccine acts, so the temporal dynamics of people vaccinated in each age group, including the times at which younger age classes are allowed, will be scenario-specific (individuals that become infected will not be vaccinated).

The vaccination scenarios have been generated by a vaccination calculator, developed for a practical use by vaccination coordinators in the Czech Republic regions (*20*). This calculator plans day-to-day vaccination for each of the assessed groups. Also, it calculates the maximum amount of applied doses by an inter-temporal choice algorithm together with several boundaries. The boundaries were calibrated by empirical data on the actual age-group vaccination in Czechia in between December 27, 2020 (start of vaccination) and March 15, 2021, with the scenarios being generated up to July 4, 2021. All data up to March 15, 2021 are therefore matching the reality. The maximal amount of daily administered doses has been set to 1*/*12 of the available doses at the respective day to account for empirically observed limits of the system. The size of group of (health care) workers was set to 550, 000, the rest of the population is vaccinated starting from the oldest ones. Each of the two groups were given 50% of each day’s capacity. For the people who were on a waiting for the second dose by the date of data generation (March 15, 2021) was the second dose date shifted according to the currently evaluated delay. The amounts of doses, delivered to particular dates, were set according to expected delivery as communicated within governments’ strategical documents and the media. See the calculator excel file attached to the reference (*20*) for a list of all sources on dose quantity.

### Vaccine effects and their timing

Given diverse and steadily appearing reports on how the vaccine may act and to what degree of efficacy, e.g., from the UK (*15, 16*) or Israel (*19*), we consider several possibilities: (1) Leaky transmission effect. Vaccinated individuals have reduced chance of infection transmission upon contact with an infectious person, such that the probability to get infected upon such a contact is reduced by the factor 1 *− v*_*e*_, where *v*_*e*_ is vaccine efficacy. Infected vaccinated individuals have the same further infection course as the non-vaccinated ones, and those that are not infected by an actual contact may get so upon further such contact. (2) All-or-nothing transmission effect. Vaccinated individuals have reduced chance to get infected upon contact with an infectious person, such that after two weeks following the first dose, a vaccinated individual becomes 100% protected with probability *v*_*e*_ and 0% protected with probability 1 *− v*_*e*_. Individuals that do not become protected after the first dose may become so a week after the second dose, with probability of 100% protection corresponding to the difference between second dose and first dose efficacies. (3) Probability of becoming symptomatic. Vaccine does not affect chance of infection transmission, but when a vaccinated individual gets infected, probability of getting symptoms is reduced by the factor 1 *− v*_*e*_. (4) Variants 1 and 3 combined. (5) When a vaccinated individual gets infected and shows symptoms, probability of becoming hospitalized is reduced by the factor 1 *− v*_*e*_. (6) Variants 1, 3 and 5 combined. (7) Variant 6, extended to account also for the action on the probability of needing an ICU and on the probability of dying when on ICU, both reduced by the factor 1 *− v*_*e*_.

### Model H

One of the models we consider is a deterministic compartmental model the major focus of which is on dynamics of hospitalizations. Because of that, symptomatic individuals may have either mild symptoms and be isolated at home, or have severe symptoms and get hospitalized. Hospitalized individuals are first put on a common bed, with some of them later getting worse and moving to an ICU, where after some time a proportion of individuals die. Individuals that do not go to the ICU recover, and those that improve on the ICU return to a common bed for a while to eventually recover and leave hospital. This model discerns four age cohorts: 0-19 years (children), 20-64 years (adults), 65-79 years (seniors), and 80+ years (elderly).

Vaccination is here implemented as follows. Only susceptible individuals are vaccinated, and three sequential vaccination classes are considered. Individuals just getting the first vaccine dose go to the first of these classes that corresponds to no vaccine effect for the first two weeks after the first dose. If not infected during this period, they pass to the second class where they stay for until one week after the second dose is administered; vaccine efficacy 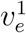 is associated with this class. Finally, if still not infected, they eventually pass to the third class for which vaccine efficacy is 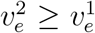. Individuals that are infected in any vaccine class stay in that class and go through the analogous sequence of events as the infected non-vaccinated individuals.

This model is calibrated on real time series of the actual number of hospitalized individuals and the total number of COVID-19-related death, using the stochastic Approximate Bayesian Computation (ABC) technique (*21,22*). More details on this model, including equations, model parameters and specific values used to run it are provided in the Electronic Supplementary Material. In particular, the calibration period is from August 31, 2020, until February 15, 2021. Within this period, epidemic dynamics was largely modulated by temporal changes in the number of social contacts, surveyed throughout all waves of COVID-19 in Czechia (*23*); Fig. 7. Then, until March 1, 2021, contacts were set at 55%, respecting results of further rounds of that panel survey. Since March 1, 2021, further restriction were imposed in Czechia, with sociological data suggesting contacts at 45%. We keep this value until present, even though some intervention relaxation has already started. We also account for the B.1.1.7 variant of SARS-CoV-2 virus in this models, where we implement its effect as a linear increase in infection transmission by 50% from January 1, 2021, to March 1, 2021. Last but not least, for validation purposes, we assume three weeks difference between the first and second vaccine doses, with 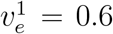 and 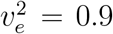 and the action on leaky infection transmission. A sound fit has been achieved for the model, at the level of both total and age-specific populations (Fig. 7).

**Figure 7:**
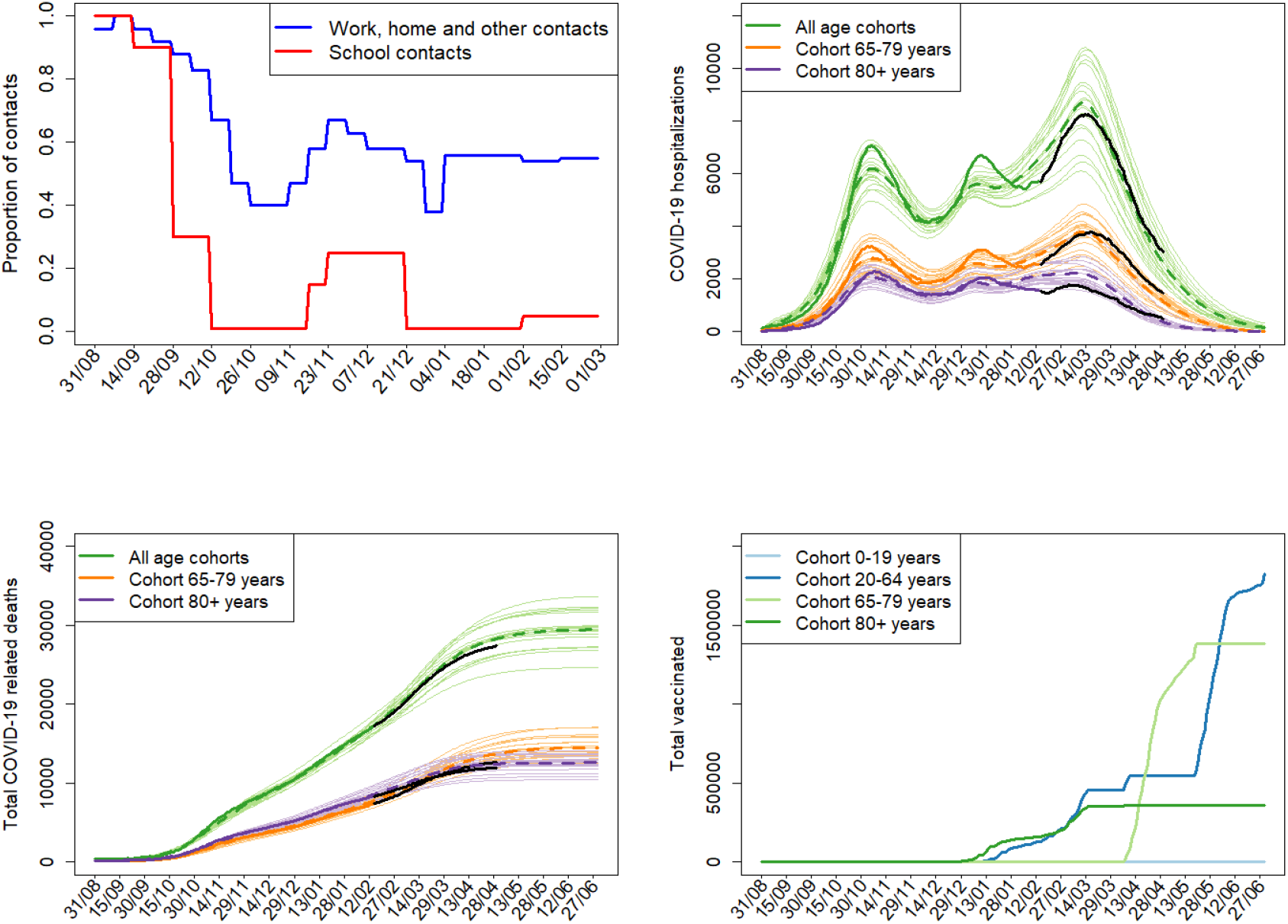
Model H calibration results performed via the Approximate Bayesian Computation technique. The top left panel: proportional reduction of contacts due to COVID-19-related interventions in Czechia; since March 1, 2021, contacts are set at 45% outside schools and 1% in schools. The top right and bottom left panels: while thin lines correspond to the best 20 worlds out of 100.000 simulated ones (selected on the basis of the smallest Euclidean distance between simulated and observed data), thick dashed lines are respective mean model trajectories. While the thick solid lines are real data up to February 15, 2021, that is, data used for calibration, the black lines are data after that date, providing sound model validation. The bottom right panel: daily numbers of the first vaccine doses administered among the considered age cohorts with 21-day delay.

### Model M

An alternative model used in the experiments is an agent-based model with agents representing a synthetic population of 56,000 people connected by a realistic network of social contacts. This network comprises 2.7 million edges in 30 layers corresponding to various types of contact, from families and neighborhood to work, school and public transportation. The population and its contact network represent the Hodonin county in the Czech Republic. The underlying epidemic model that runs for every agent is a SEIR-type model with asymptomatic and presymptomatic infectious classes, and a parallel set of states for individuals detected through testing and contact tracing. The model allows for individually assigned parameters based on age, sex, level of protection and other characteristics.

The vaccination is here implemented as a special policy with several parameters operating on the individual level. Non-detected individuals are vaccinated in a stochastic manner according to the given scenario which reflects (age-based) vaccination constrains and preferences. The vaccinated node is marked as vaccinated and has a new counter: the number of days since first vaccine dose. As soon as this counter reaches 14 days, the vaccine efficacy 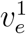 is applied. And, as the counter reaches δ + 7 days (where δ is the delay between the first and second vaccine shot), actions are implemented with vaccine efficacy 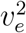.

This model is fitted to real time series on COVID-19-related deaths. The calibration was done using extensive grid search. More details about the underlying model and its parameters can be found in (*24*). In particular, the model was calibrated on data from October 5, 2021, until February 17, 2021 (Fig. 8). The contact rates in 30 layers were set according to contact reduction in the Czech public given in (*23*) and were kept constant from January 24, 2021.

**Figure 8:**
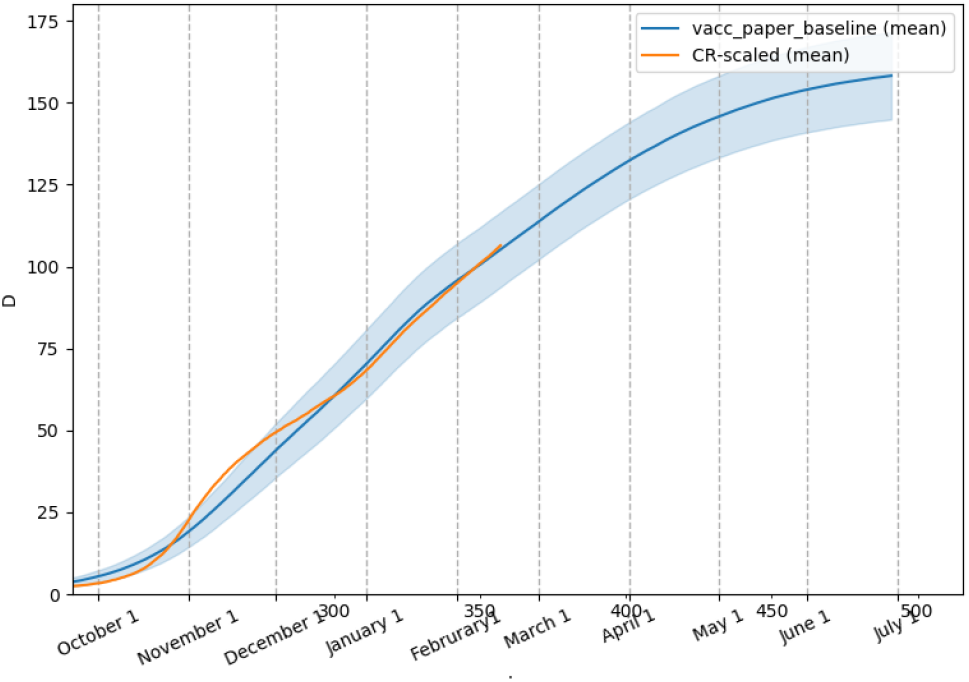
Calibration of Model M. The *y*-axis shows the model-predicted cumulative number of COVID-19-related deaths in simulation (blue, mean *±* one standard deviation) compared to the real situation in the Czech Republic scaled to the model population size (orange).

### Model F

Our third model is a stochastic, discrete-time SEIR model distinguishing the same four age cohorts as Model H. This model takes into account both the visible part of the epidemic (i.e. cases revealed by testing) and its hidden part (a certain proportion of asymptomatic and symptomatic cases remain undetected). The course of epidemic is modeled as dependent on the social contact restrictions and the level of fear from epidemic, both surveyed by (*23*); thus, the simulated vaccination scenarios are realistic in the sense that they use true values of these determinants.

The vaccination is implemented as follows. First, the probability 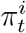 of being protected within the *i*-th age cohort at time *t* is determined as 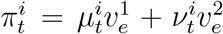, where 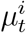 and 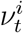 are the ratios of individuals two weeks after the first dose and one week after the second dose, respectively, and 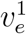 and 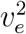 are the corresponding efficacies. For the leaky transmission approach, the force of infection (transition rate from *S* to *E* classes) is simply multiplied by 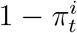. For the all-or-nothing transmission approach, in addition to this reduction, the transition from *S* to *R* classes is considered with the rate 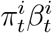, where 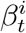 is the original infection rate. Finally, for the “symptoms reduction” variant, the infection rate is left intact yet the probability that a vaccinated person becomes symptomatic after getting infected is multiplied by 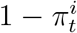.

The model is calibrated by real time series on reported cases, admissions and releases from hospitals, deaths due to COVID-19, and the overall rate of asymptomatic cases revealed by testing and contact tracing. For details, see (*25*). In particular, data for calibration cover the period up to April 10, 2021, after which we assumed contact reduction at 50%. For the calibration, we assumed (and estimated) different infection transmission and hospitalization rates of the B.1.1.7 variant of the SARS-CoV-2 virus, gradually prevailing since January until March in the Czech Republic. Using vaccine efficacies 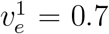 and 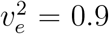 of the first and second doses, respectively, calibration and validation results are provided in Fig. 9.

**Figure 9:**
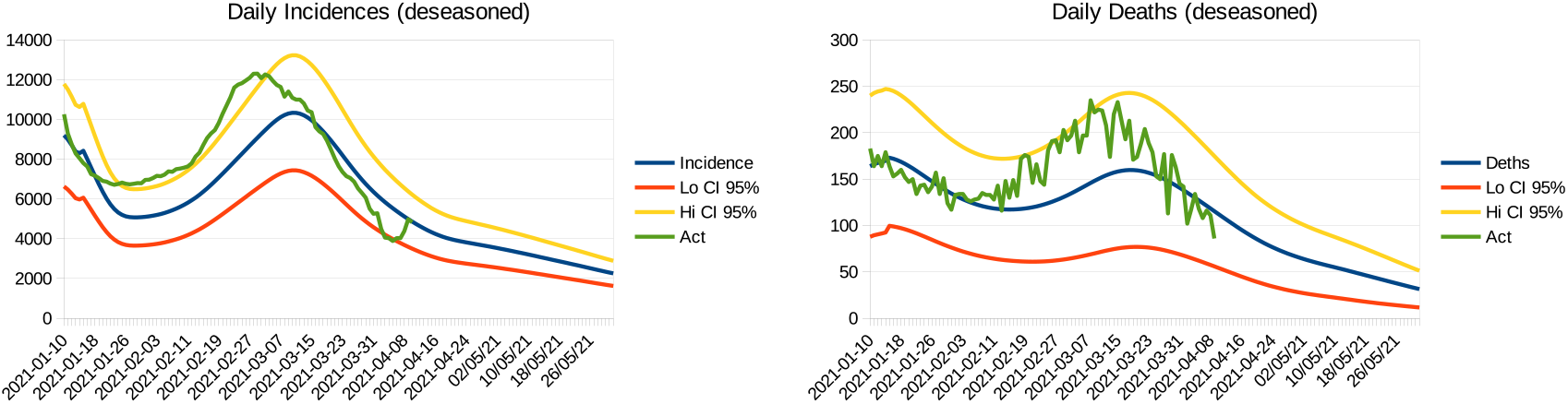
Model F calibration. Left panel: daily detected incidences of COVID-19. Right panel: daily deaths due to COVID-19. Legend: blue line is the mean prediction, while the yellow and red lines delimit the 95% confidence interval. Green curve represents real data.

## Data Availability

Computer codes of all involved models are available upon a justified request.

## Acknowledgments

The authors thank The Institute of Health Information and Statistics of the Czech Republic (UZIS) for providing us detailed data on many characteristics of the COVID-19 epidemic in Czechia. We also acknowledge the PAQ Reseach agency for sharing with us the results from their sociological surveys.

## Author Contributions

LB, RL, MS and RN conceived the study. RL and JW prepared the vaccination scenarios. LB, MS, RN, PV and GS coded the model and performed the simulations. All authors wrote and revised the manuscript.

## Data and materials availability

Codes of models allowing to reproduce all simulations is available from the authors upon request (will be made available in a code storage place when the manuscript is accepted).

## Supplementary materials

### Materials and Methods

All three models we use to address our research question are of the SEIR type, the generally accepted framework for modeling COVID-19, and all explicitly account for asymptomatic and presymptomatic infectious classes. That is, once infected, individuals are not infectious for a period. Once infectious, they either stay asymptomatic for the rest of infection (asymptomatic state) or are asymptomatic only for a short period (presymptomatic state) before symptoms eventually appear (symptomatic state). Symptomatic individuals may have only mild symptoms in which case they are (largely) isolated at home, or may have severe symptoms requiring hospitalization. Some hospitalized individuals eventually die. In addition, all three models are structured by age of individuals and type of inter-individual contacts. On the other hand, each model has a number of unique assumptions and characteristics. Here we describe Model H that focuses on dynamics in hospitals, while referring to publicly available descriptions of Model M (*24*) and Model F (*25*).

#### Description of Model H

##### Epidemic model

Due to contacts with infectious individuals, susceptible individuals (class *S*) may become exposed (*E*), that is, infected but not yet infectious, with probability *λ* corresponding to the force of infection. The exposed individuals then become asymptomatic for the whole course of infection (*A*, with probability 1 *− p*_*S*_) or presymptomatic for just a short period of time before becoming symptomatic (*P*, with probability *p*_*S*_). Average lengths of exposed, asymptomatic and presymptomatic periods are *d*_*E*_, *d*_*A*_ and *d*_*P*_ days, respectively. The *P* individuals then become symptomatic (*I*), reducing their contacts with others by a factor *r*_*C*_ (imperfect social isolation). The *A* individuals eventually recover (*R*).

After an average period of *d*_*I*_ days, a proportion *p*_*H*_ of symptomatic individuals (those with relatively severe symptoms) are hospitalized (*H*). The remaining proportion 1 *− p*_*H*_ of symptomatic individuals (those that have only mild symptoms) remain isolated at home, staying in class *I*_*R*_ until recovery, with contacts again reduced by *r*_*C*_, which lasts on average *d*_*A*_ *− d*_*P*_ *− d*_*I*_ days. Here we assume that the average period from infection to recovery is the same for individuals with no or mild symptoms. And since the period from appearance of symptoms to recovery differs from that from symptoms occurrence to hospitalization, and the probability of hospitalization *p*_*H*_ is independent of these periods, we need to introduce an artificial inter-class *I*_*R*_.

Hospitalized individuals follow two different pathways. After *d*_*HJ*_ days spent on a common hospital bed, a proportion *p*_*J*_ of such individuals require intensive care (*J*). Individuals that do not need intensive care go after those *d*_*HJ*_ days to another inter-class *H*_*R*_ where they eventually recover after next *d*_*HR*_ days. While on an ICU, individuals may die (*D*) with probability *p*_*D*_ (after *d*_*JD*_ days). If not dying, which happens with the complementary probability 1 *− p*_*D*_, they eventually recover, after spending further *d*_*JH*_ days on the ICU (inter-class *J*_*H*_) and other *d*_*HR*2_ days on the common bed (*H*_*R*2_). All model variables are summarized in Table S1.

**Table S1:**
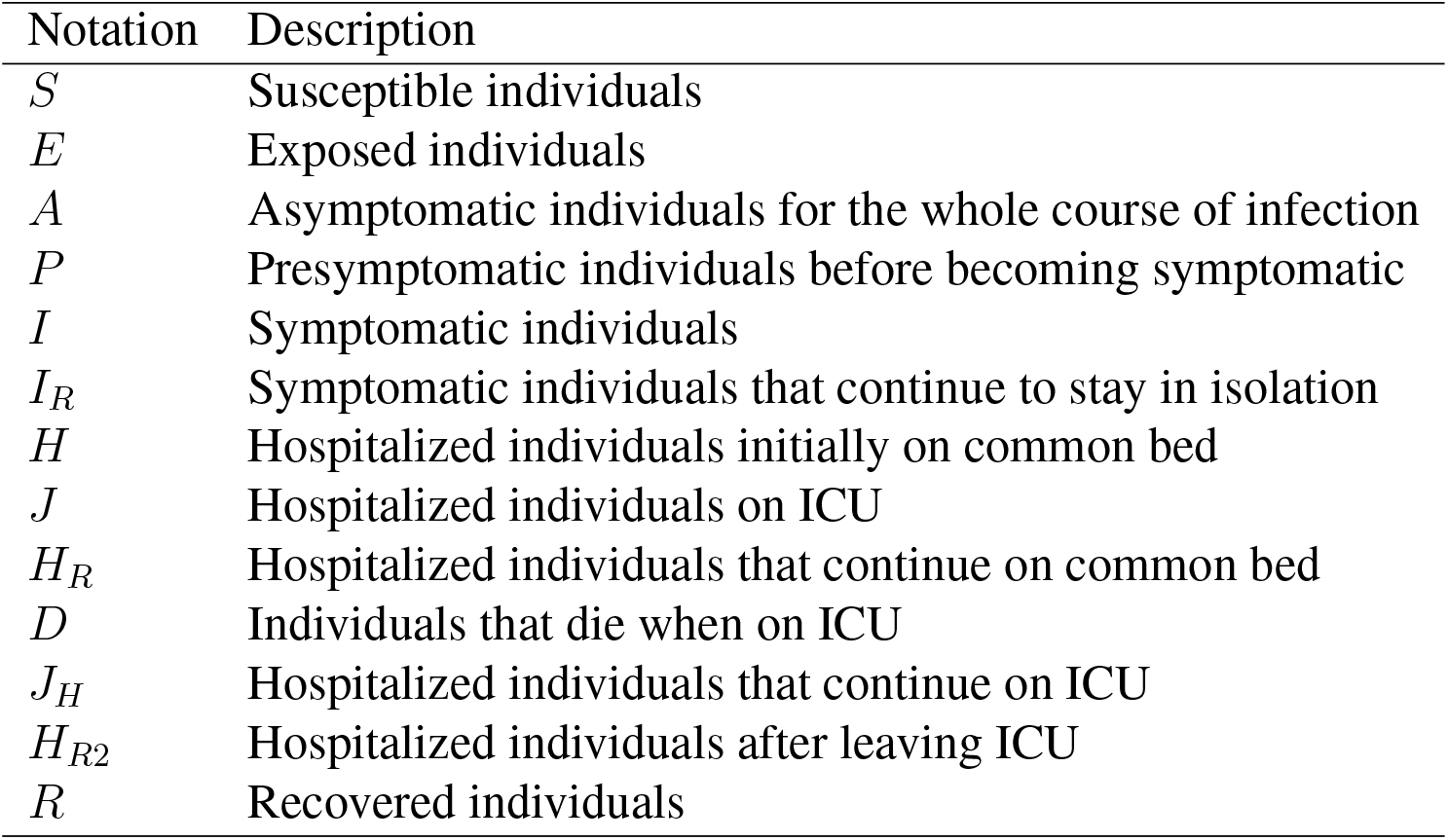
List of variables used in Model H.

| Notation | Description |
| --- | --- |
| $S$ | Susceptible individuals |
| $E$ | Exposed individuals |
| $A$ | Asymptomatic individuals for the whole course of infection |
| $P$ | Presymptomatic individuals before becoming symptomatic |
| $I$ | Symptomatic individuals |
| $I_R$ | Symptomatic individuals that continue to stay in isolation |
| $H$ | Hospitalized individuals initially on common bed |
| $J$ | Hospitalized individuals on ICU |
| $H_R$ | Hospitalized individuals that continue on common bed |
| $D$ | Individuals that die when on ICU |
| $J_H$ | Hospitalized individuals that continue on ICU |
| $H_{R2}$ | Hospitalized individuals after leaving ICU |
| $R$ | Recovered individuals |

Finally, the SARS-CoV-2 virus is known to differently impact various age cohorts (*27*). Therefore, we distinguish four age cohorts, 0-19 years (named children, coded 1), 20-64 years (adults, 2), 65-79 years (seniors, 3), and 80+ years (elderly, 4). These classes interact via the force of infection (see below). Once infected, individuals of each age cohort proceed independently of individuals of other age cohorts. Nonetheless, some model parameters used to decide on specific pathways through the model are age-specific. These are probabilities of becoming symptomatic *p*_*S*_, requiring hospitalization *p*_*H*_, needing ICU *p*_*J*_, and dying *p*_*D*_, but also all periods of staying in various hospital classes.

In discrete time, with one time step corresponding to one day, the above model description can be translated into the following system of equations:

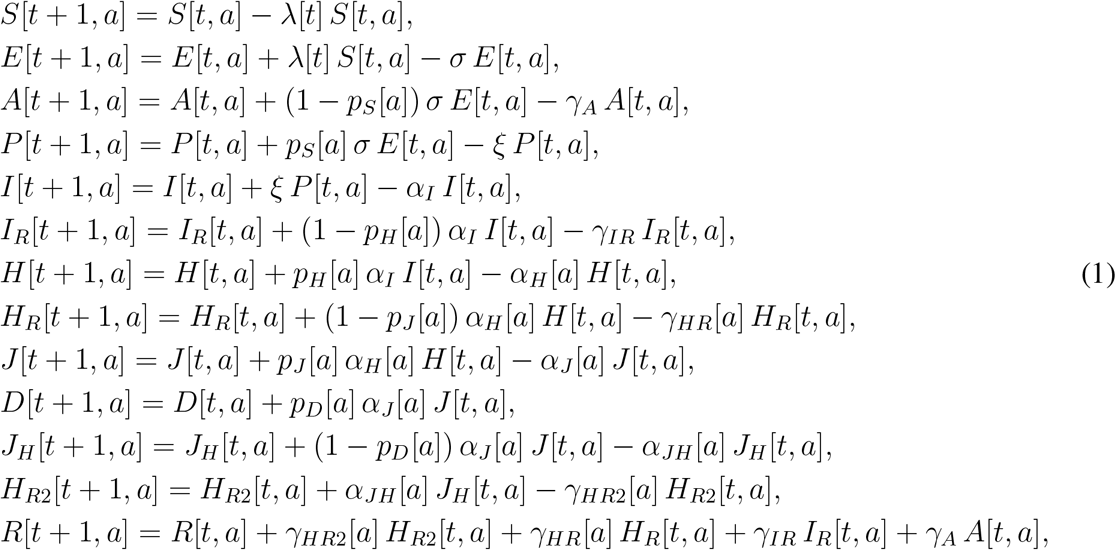

where all variables are functions of time *t* and age cohort *a* = 1, 2, 3, 4. The model parameters *σ, ξ, α*_*I/H/J/JH*_, and *γ*_*A/IR/HR/HR*2_ represent probabilities with which individuals leave respective model classes. These probabilities are related to the average periods an individual spends in each such class (Table S2).

**Table S2:** Probabilities of leaving particular model classes calculated from average periods spent in those classes.

| Notation | Description | Relationship to delays |
| --- | --- | --- |
| $\sigma$ | Probability of leaving $E$ class | $1 - \exp(-1/d_E)$ |
| $\xi$ | Probability of leaving $P$ class | $1 - \exp(-1/d_P)$ |
| $\alpha_I$ | Probability of leaving $I$ class for $H$ or $I_R$ classes | $1 - \exp(-1/d_{IH})$ |
| $\alpha_H$ | Probability of leaving $H$ class for $J$ or $H_R$ classes | $1 - \exp(-1/d_{HJ})$ |
| $\alpha_J$ | Probability of leaving $J$ class for $D$ or $J_H$ classes | $1 - \exp(-1/d_{JD})$ |
| $\alpha_{JH}$ | Probability of leaving class $J_H$ for class $H_{R2}$ | $1 - \exp(-1/d_{JH})$ |
| $\gamma_A$ | Probability of leaving class $A$ for recovery | $1 - \exp(-1/d_A)$ |
| $\gamma_{IR}$ | Probability of leaving class $I_R$ for recovery | $1 - \exp(-1/d_{IR})$ |
| $\gamma_{HR}$ | Probability of leaving class $H_R$ for recovery | $1 - \exp(-1/d_{HR})$ |
| $\gamma_{HR2}$ | Probability of leaving class $H_{R2}$ for recovery | $1 - \exp(-1/d_{HR2})$ |

##### Force of infection

The force of infection *λ* in the model (1) sums contributions from all infectious classes (*A, P, I, I*_*R*_) across all age cohorts we consider:

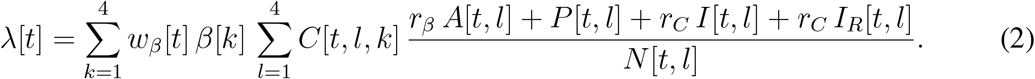

Here, *β*[*k*] is the age-specific probability that a susceptible individual from age cohort *k* is infected by a (sufficiently close and lengthy) with an infectious individual of any age cohort, *C*[*t, l, k*] is the contact rate (the mean number of individuals of age *l* that an individual of age *k* has an effective contact with during day *t*), *r*_*β*_ is a factor reducing the infection transmission probability for an asymptomatic individual relative to a (pre)symptomatic, *r*_*C*_ is a factor reducing the contact rate of a symptomatic individual relative to an a/presymptomatic one (having symptoms should force an individual to reduce contacts with others but in reality isolation is imperfect), and *N* [*t, l*] is the number of ‘active’ age *l* individuals at time *t*, that is, those except dead, in hospitals and symptomatic individuals that are isolated. All parameters and their values we run the model with are given in Table S3.

In addition, *w*_*β*_[*t*] is the time-varying (but age-independent) factor reducing (in case of personal protective measures such as masks) or enhancing (in case of more contagious viral variants) the probability of infection transmission upon contact *β*. We may thus view it as a product of reducing and enhancing forces. We set the reducing force to 0.3 until February 28, 2020, and decrease it further to 0.23 from March 1, 2021, the day from which FN95 respirators were ordered to wear instead of common medical masks. We calculate these numbers as

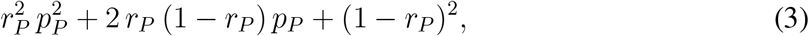

where *r*_*P*_ = 0.6 represents 60% compliance of using personal protection measures and *p*_*P*_ is the proportional reduction in transmission due to personal protection measures. We set *p*_*P*_ to 0.25 until February 28, 2020, and to 0.125 from March 1, 2021. Moreover, we increase the enhancing force from 1 to 1.5 since January 1, 2021, to March 1, 2021, to account for invasion and eventual domination of the B.1.1.7 variant of SARS-CoV-2 virus in the Czech Republic.

Finally, construction of the time-varying contact matrix *C* starts with (*26*), a study in which such a matrix was published for 152 countries, including Czechia, for the pre-pandemic times. They expressed it as a sum of four specific contact matrices describing daily numbers of contacts at home (*C*_*H*_), school (*C*_*S*_), work (*C*_*W*_), and of other types of contact (*C*_*O*_). We transform the Czech matrices of (*26*), structured by 5-year age classes, to fit our three age cohorts. This is because data in (*26*) end with the 75-80 years age class. To supply values for our age cohort 4 (adults 80+ years), we replicate data for our age cohort 3 (adults 65-79 years) to our age cohort 4, too, to get:

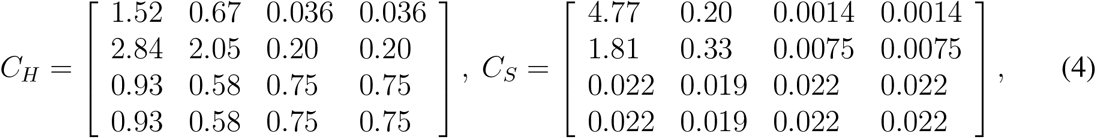

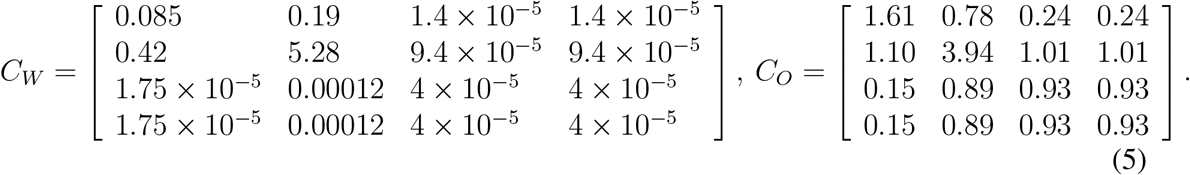

The time-varying contact matrix *C* is then calculate as a weighted sum of these four specific matrices,

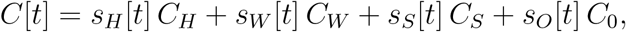

where the weights *s*_*X*_, *X* = *H, W, S, O* are time-varying (changing on a weekly basis) proportional reduction factors due to adopted interventions and their compliance, estimated from panel surveys conducted by the PAQ Reseach agency^1^ (Fig.S1).

**Figure S1:**
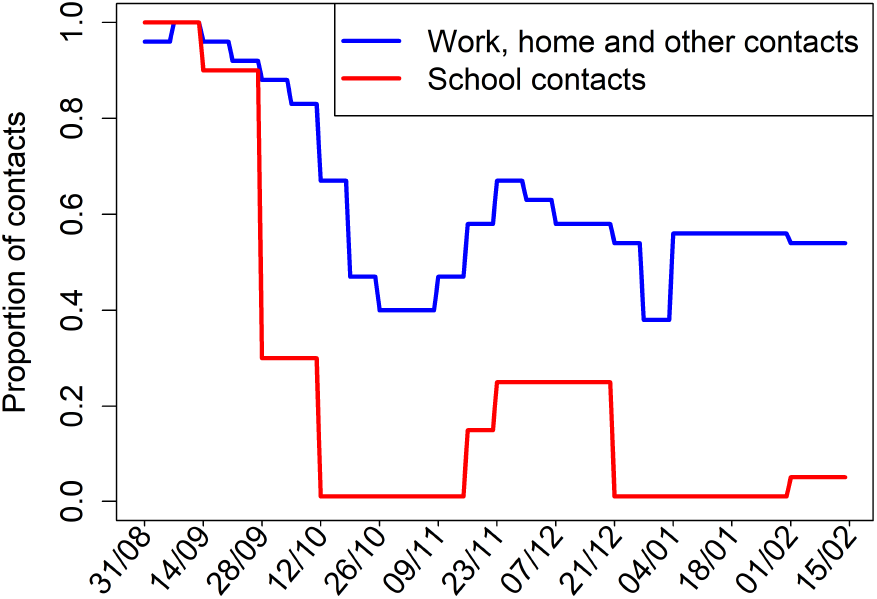
Proportional reductions in different types of contact evolving in time on a weekly basis. Based on panel surveys conducted by the PAQ Reseach agency (www.zivotbehempandemie.cz).

##### Initial conditions

Our simulations start on August 31, 2020, the day around which the second COVID-19 wave in Czechia presumably began and also corresponds to Monday (contact data described above are provided for calendar weeks). Whereas initial values of state variables related to hospitals are inferred from data, initial values for the remaining (hidden) state variables (*E, A, P, I, I*_*R*_) are regarded as parameters to which the model is calibrated; see below for the calibration procedure.

##### Vaccination

Only susceptible individuals are vaccinated. When vaccinated, individuals pass through three sequential vaccination classes. Just vaccinated individuals enter the class with no vaccine effect in which they stay on average for two weeks. If not infected during this time, they pass to the second class for until one week after the second dose; the vaccine efficacy is 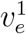 when in this class. Eventually, if still not infected, vaccinated individuals pass to the third class in which vaccine efficacy is 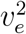. Vaccinated individuals can be infected when in any of these three classes. Once this happens, they stay in the current class and go through the sequence of states analogous to that for non-vaccinated infected individuals.

With these three classes of vaccinated individuals, the force of infection changes to

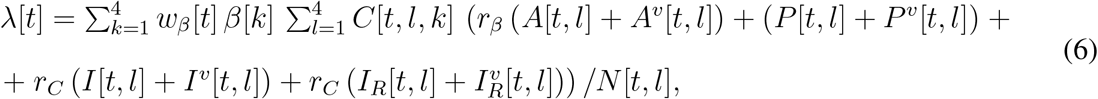

where we assume that the infected vaccinated individuals are infectious to the same extent as the infected non-vaccinated individuals, and where the terms with the upper index *v* are numbers of the respective individuals over all three vaccination classes.

The ways vaccine is in this study assumed to acts in four ways (and some of their combination). These ways and the manner in which they are modeled are as follows:

1. Reduction of the probability of getting infected upon contact with an infectious individual (the ‘leaky’ variant): the force of infection *λ*[*t*] is reduced by the factor 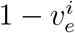;
2. Reduction of the probability of becoming symptomatic when infected: the probability *p*_*S*_ is reduced by the factor 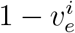;
3. Reduction of the probability of getting hospitalized when symptomatic: the probability *p*_*H*_ is reduced by the factor 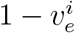;
4. Reduction of the probability of getting infected upon contact with an infectious individual (the ‘all-or-nothing’ variant): two weeks after the first dose, the vaccinated individual is fully protected with probability 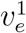 and completely unprotected with complementary probability; when unprotected, then after the second dose it becomes fully protected with probability 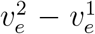, while it remains completely unprotected with the complementary probability;
5. Reduction of the probability of needing ICU when getting hospitalized: the probability *p*_*J*_ is reduced by the factor 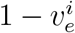;
6. Reduction of the probability of dying when on ICU: the probability *p*_*D*_ is reduced by the factor 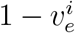.

##### Model calibration

Many model parameters can be taken from the literature or estimated from data provided by the Institute of Health Information and Statistics of the Czech Republic (UZIS); www.uzis.cz. However, values of some model parameters will always remain uncertain, of which the transmission probabilities *β*[*i*], *i* = 1, 2, 3, 4, are commonly of this kind. These and some other model parameters, listed in Table S3, are estimated by fitting the simulated cumulative numbers of deaths and the actual numbers of hospitalized individuals in age classes 2, 3 and 4 to the corresponding age-specific actual time series collected in the Czech Republic.

Many optimization and filtering methods have been developed to meaningfully perform model calibration (*30*). Here we adopted the Approximate Bayesian Computation (ABC) technique, used to estimate parameters of complex models in genomics and other biological disciplines, including epidemiology (*21, 22, 32, 34, 35*). The major advantage of this method is that it naturally works with all sources of uncertainty acknowledged in the model. At the same time, the ABC does not rely on likelihood calculation and in case of sufficient computation power can be used with models of virtually any complexity.

The variant of ABC with rejection sampling that we used consisted of three steps. First, we performed *K* = 200, 000 model simulations, collecting the age-specific cumulative numbers of deaths (*D*) and actual numbers of hospitalized individuals (*H* + *H*_*R*_ + *J* + *J*_*H*_ + *H*_*R*2_), drawing values of the uncertain model parameters from prior distributions based on literature and available data on the Czech Republic epidemic; selected prior distributions for the parameters to be estimated are given in Table S3. Second, we calculated summary statistics on the simulated and corresponding observed time series, using their Euclidean distance *D*. Third, we selected model simulations that satisfied *D < ϵ*, where *ϵ* was chosen to pass 0.025% (50) of the simulations into the selected set. Since the used summary statistics are informative, the distribution of parameters corresponding to the selected simulations is known to converge from outside to the Bayesian posterior distribution of parameter values with *N* going to infinity and *ϵ* going to 0, and is referred to as the approximate posterior (*32*). The choice of *N* and *ϵ* in the ABC is driven by compromise between computation power and smoothness and accuracy of the approximate posterior.

The set of selected parametric sets thus allows us to evaluate remaining parameter uncertainty, given the available data and adopted summary statistics (*21, 22, 32*). This is crucial to realize, since although different parameter sets may similarly fit the available data (have similar summary statistics), and often provide similar short-term predictions, they may demonstrate significant differences in longer term and in interplay with intervention policies. To apply the ABC technique, we use the `abc` package in R (*36*), modified to work with non-normalized summary statistics.

**Table S3:** Parameters of Model H. UZIS = The Institute of Health Information and Statistics of the Czech Republic (www.uzis.cz); mean values are provided over the period from August 31, 2020, until February 14, 2021. Distributions of all the indicated delays were checked and all are exponential. Times are given in days. The indicated distributions are prior distributions for the calibration procedure, and are based on the literature.

| Parameter | Meaning | Value | Reference |
| --- | --- | --- | --- |
| $\beta_1, \beta_2, \beta_3, \beta_4$ | Age-specific transmission probabilities | uniform on $[0.01, 0.99]$ | calibration |
| $r_\beta$ | Factor reducing infection transmission from asymptomatic individuals | 0.5 | (27) |
| $r_C$ | proportional contact reduction in isolated individuals | uniform on $[0.01, 0.4]$ | calibration |
| $p_S$ | proportion symptomatic | uniform on $[0.65, 0.84]$ | calibration, halved for the 0-19 years age cohort |
| $p_H$ | proportion hospitalized among symptomatic | $(0.0052, 0.056, 0.270.43)$ | UZIS |
| $p_J$ | proportion on ICU among hospitalized | uniform on $(0 - 0.02, 0 - 0.3, 0.1 - 0.4, 0.1 - 0.7)$ | UZIS<br>$(0, 0.16, 0.24, 0.19)$, calibration |
| $p_D$ | proportion dying on ICU | uniform on $(0 - 0.02, 0.1 - 0.5, 0.3 - 0.9, 0.5 - 0.9)$ | UZIS<br>$(0, 0.24, 0.51, 0.74)$, calibration |
| $d_E$ | latent period | uniform on 3-7 days | calibration |
| $d_A$ | time to recovery when asymptomatic | uniform on 8-14 days | calibration |
| $d_P$ | presymptomatic period | uniform on 2-6 days | calibration |
| $d_{IH}$ | period from symptoms appearance | $(5.71, 7.41, 5.87, 5.87)$ days | UZIS |
| $d_{IR}$ | recovery time when isolated | $d_A - d_P - d_{IH}$ | – |
| $d_{HJ}$ | period from hospital to ICU admission | $(1.88, 3.01, 3.85, 4.50)$ | UZIS |
| $d_{JD}$ | time on ICU till death | $(10.0, 9.41, 7.89, 4.43)$ | UZIS |
| $d_{JH}$ | extra time on ICU when not dying | $(0, 0, 0.71, 2.22)$ | UZIS |
| $d_{HR2}$ | time from leaving ICU to recovery | $(3.08, 4.51, 4.95, 6.12)$ | UZIS |

## Footnotes

1 www.zivotbehempandemie.cz

## References and notes

1. G. Giordano, et al., Nature Medicine (2021).

2. S. Moore, E. M. Hill, M. J. Tildesley, L. Dyson, M. J. Keeling, Lancet Infectious Diseases (2021).

3. M. Amaku, D. T. Covas, F. A. B. Coutinho, R. S. Azevedo, E. Massad, medRxiv (2021).

4. A. D. Paltiel, J. L. Schwartz, A. Zheng, R. P. Walensky, Health Affairs 40, 42 (2021).

5. Fact sheet for healthcare providers administering vaccine (vaccination providers), www.fda.gov/media/146304/download. Accessed: 2021-06-09.

6. Covid-19: vaccination overview in the czech republic, onemocneni-aktualne.mzcr.cz/vakcinace-cr. Accessed: 2021-06-09.

7. Interim clinical considerations for use of COVID-19 vaccines currently authorized in the United States, https://www.cdc.gov/vaccines/covid-19/info-by-product/clinical-considerations.html. Accessed: 2021-06-09.

8. Specific situations regarding vaccination in the czech republic, covid.gov.cz/situace/informace-o-vakcine/specificke-situace-pri-ockovani. Accessed: 2021-06-09.

9. K. M. Bubar, et al., Science 371, 916 (2021).

10. S. Moore, E. M. Hill, L. Dyson, M. J. Tildesley, M. J. Keeling, PLoS Computational Biology 17(5), e1008849 (2021).

11. A. D. Paltiel, A. Zheng, J. L. Schwartz, Annals of Internal Medicine (2021).

12. A. R. Tuite, D. N. Fisman, L. Zhu, J. A. Salomon, Annals of Internal Medicine (2021).

13. Delaying second Pfizer vaccines to 12 weeks significantly increases antibody responses in older people, finds study, https://www.birmingham.ac.uk/news/latest/2021/05/covid-pfizer-vaccination-interval-antibody-response.aspx. Accessed: 2021-06-09.

14. F. P. Polack, et al., New England Journal of Medicine 383, 2603 (2020).

15. E. Vasileiou, et al., SSRN (2021).

16. V. J. Hall, et al., Lancet 397, 1725 (2021).

17. D. M. Skowronski, G. D. Serres, New England Journal of Medicine 384, 1577 (2021).

18. N. Dagan, et al., New England Journal of Medicine 384, 1412 (2021).

19. E. J. Haas, et al., Lancet (2021).

20. J. Weiner, E. Blechová, R. Levinský, R. Horká, Do kdy je možné naočkovat nejrizikovější skupiny? (2021).

21. T. Toni, D. Welch, N. Strelkowa, A. Ipsen, M. P. H. Stumpf, Journal of The Royal Society Interface 6, 187 (2009).

22. K. Csilléry, M. G. Blum, O. E. Gaggiotti, O. François, Trends in Ecology and Evolution 25, 410 (2010).

23. P. Research, Life during pandemic (2020). Long-term sociological panel survey.

24. L. Berec, et al., medRxiv (2021).

25. M. Smid, et al., medRxiv (2021).

26. K. Prem, A. R. Cook, M. Jit, PLoS Computational Biology 13, e1005697 (2017).

27. N. G. Davies, et al., Nature Medicine 26, 1205 (2020).

## References and notes

28. N. G. Davies, et al., Nature Medicine 26, 1205 (2020).

29. K. Prem, A. R. Cook, M. Jit, PLoS Computational Biology 13, e1005697 (2017).

30. W. Yang, A. Karspeck, J. Shaman, PLoS Computational Biology 10, e1003583 (2014).

31. T. Toni, D. Welch, N. Strelkowa, A. Ipsen, M. P. H. Stumpf, Journal of The Royal Society Interface 6, 187 (2009).

32. M. A. Beaumont, Annual Review of Ecology, Evolution, and Systematics 41, 379 (2010).

33. K. Csilléry, M. G. Blum, O. E. Gaggiotti, O. François, Trends in Ecology and Evolution 25, 410 (2010).

34. M. G. Blum, V. C. Tran, Bioinformatics 11, 644 (2010).

35. F. Luciani, S. A. Sisson, H. Jiang, A. R. Francis, M. M. Tanaka, PNAS 106, 14711 (2009).

36. K. Csilléry, L. Lemaire, O. François, M. G. Blum, abc: tools for Approximate Bayesian Computation (ABC) (2015).

